# Cytokine and Lymphocyte Profiles in COVID-19 Patients with Cancer: Implications for Disease Severity and Clinical Outcomes

**DOI:** 10.1101/2025.04.24.25326371

**Authors:** Marina M. Burlá, Karina L. Silva, Bárbara C. Peixoto, Livia R. Goes, Isaclaudia Azevedo-Quintanilha, Fernando A. Bozza, Marcelo A. Soares, Andreia C. de Melo, Eugenio D. Hottz, Patricia T. Bozza, João P.B. Viola

**Affiliations:** Program of Immunology and Tumor Biology; Program of Genetics and Tumor Virology; Brazilian National Cancer Institute (INCA), Rio de Janeiro, Brazil; Division of Clinical Research and Technological Development, Brazilian National Cancer Institute (INCA), Rio de Janeiro, Brazil; Faculty of Medicine, Institute of Medical Education (IDOMED), Rio de Janeiro, Brazil; Laboratory of Immunopharmacology, Oswaldo Cruz Institute (IOC), Oswaldo Cruz Foundation (FIOCRUZ), Rio de Janeiro, Brazil; Intensive Care Medicine Laboratory, National Institute of Infectiology (INI), Oswaldo Cruz Foundation (FIOCRUZ), Rio de Janeiro, Brazil; D’Or Institute for Research and Education (IDOr), Rio de Janeiro, Brazil; Laboratory of Immunothrombosis, Department of Biochemistry, Federal University of Juiz de Fora (UFJF), Minas Gerais, Brazil

**Keywords:** Cancer, COVID-19, T lymphocyte, NK cells, Treg cells, T cell exhaustion, CXCL10/IP-10, MIF

## Abstract

Patients with cancer face a higher risk of severe complications, including death, when affected by COVID-19. Particularly in low- and middle-income countries, COVID-19 pandemic has placed a heavy burden on the public health systems, necessitating adjustments in the clinical management of cancer patients. Numerous factors have been identified to influence susceptibility to SARS-CoV-2 infection and disease severity, but the determinants of severe outcomes remain largely unknown. This study aims to characterize the cytokine and lymphocyte profiles of cancer patients with COVID-19, correlate these profiles with disease severity, and compare them to non-cancer patients. Our findings revealed reduced CXCL10 (IP-10) and MIF levels in cancer patients with COVID-19 and discriminated against disease severity. CXCL10 was further elevated in severe COVID-19 cancer patients compared to mild COVID-19 cancer patients. Additionally, cancer patients with COVID-19 exhibited reduced T lymphocytes, expansion of regulatory cells, a shift from effector memory to central memory T-cells, and increased levels of exhausted T lymphocytes. In conclusion, our data suggest that the distinct immunological profile observed in cancer patients with COVID-19 may negatively impact the clinical outcomes, highlighting potential implications for cancer patient management.

## Introduction

COVID-19 pandemic brought several issues and worries to the public health system. According to the World Health Organization (WHO), since its outbreak in 2019, this respiratory-transmitted disease occasioned more than 6 million deaths worldwide (1). Among patients with baseline diseases and comorbidities, including cancer, there is a pattern of increased susceptibility to worse prognosis (2,4).

Oncologic patients exhibit higher mortality rates and severe outcomes when compared to the general population, either because of their underlying illness pathophysiology or because of their immunosuppressive drug regimen (2,4). Therefore, since vaccinations have begun worldwide, they have been considered a priority group (4). Moreover, several studies were developed to understand their immune response to different vaccines and to evaluate baseline conditions and immunosuppressive treatments (4,5). However, given that cancer patients are still poorly contemplated in vaccination trials, there is limited knowledge regarding the degree of immunological protection guaranteed by vaccines (4). Thus, infection prevention and treatment optimization remain crucial in their management guidelines (3). Furthermore, the frequent rise of new strains and their high transmissibility are still concerns when dealing with oncologic patients, even in the post-vaccination era (3).

Studies aiming to explain the pathogenesis and immunology of COVID-19 keep being developed. According to current evidence, severe cases could be related to a depleted lymphocyte population, an exhausted T cell profile, and an increased concentration of inflammatory cytokines (7,8). Even so, the lack of clarifying studies related to the virus in cancer patients complicates optimized management, due to still unknown molecular and immunologic aspects of a patient presenting both conditions at the same period, which further damages the evaluation of their clinical outcomes. Increased PD-1, CD95, and the expression of apoptotic molecules have been previously studied as possible biomarkers of COVID-19 severeness in cancer patients (8). However, these studies highlight the validation difficulty of the findings, implying that future research is necessary for further understanding the mechanisms involved in this pathology (8). Therefore, this study aimed to evaluate the cytokine and cellular activation profile of cancer patients with COVID-19 and to correlate it with their outcome severeness, including death and mechanical ventilator use.

Moreover, in this study we aim to compare the cytokine and lymphocyte profiles of COVID-19 cancer patients with non-cancer patients, targeting to correlate a specific profile and their disease severity. Our data showed that CXCL10 (IP-10) and MIF were differently produced in COVID-19 patients with cancer and discriminated disease severity. Severe COVID-19 cancer patients show reduced T lymphocytes, an amplification of regulatory lymphocytes (T and NK cells) and an exhausted immune profile that could have a potential negative implication for cancer clinical outcomes.

## Methods

### Study design and participants

This is a prospective cohort study of cancer patients with mild or severe COVID-19 and non-cancer patients with mild or severe COVID-19. Cancer patients were selected from electronic medical records and compiled data of inpatients admitted to INCA (Instituto Nacional de Câncer, Rio de Janeiro, Brazil) from June 2020 to August 2020. Hospital admissions occurred for COVID-19 symptoms or other medical reasons. Contact with positive COVID-19 patients and symptoms throughout hospitalization were also reported. Non-cancer patients with laboratory-confirmed SARS-CoV-2 with COVID-19 within 72 hours from ICU admission in three reference centers (Instituto Estadual do Cérebro Paulo Niemeyer, Hospital Copa Star and Leblon Campaign Hospital, all in Rio de Janeiro, Brazil) from April 2020 to August 2020. Severe COVID-19 was defined as those critically ill patients, presenting viral pneumonia on computed tomography scan and requiring oxygen supplementation through either a nonrebreather mask or mechanical ventilation.

The diagnosis and stratification of COVID-19 patients were based according to the WHO guidelines. Diagnostic confirmation was defined by a positive result on real-time reverse transcriptase polymerase chain reaction (RT-PCR) assay of nasal- and oropharyngeal swab specimens using the U.S. Centers for Disease Control and Prevention (CDC) reagents and protocol (9).

This study was approved by the Brazilian National Commission of Ethics in Research (approval number: CAAE 30608220.8.0000.5274 and CAAE 30650420.4.1001.0008) and conducted following the Good Clinical Practice guidelines, keeping participant identities confidential.

### Data collection and analysis

Demographic and clinical features, such as comorbidities, tumor subtype, stage, metastatic sites, cause of death, and laboratory tests at diagnosis during hospitalization were collected from the medical records. Clinical treatments for cancer and COVID-19 were also collected. Patients were classified as severe COVID-19 in cases requiring oxygen supplementation through either a nonrebreather mask or mechanical ventilation and/or death caused by the infection. Otherwise, patients were classified as mild COVID-19 cases. Patients who had not been discharged from the hospital were censored for 28-day mortality.

We collected plasma from, COVID-19 patients with or without cancer to evaluate their immunological profiles. We analyzed 42 different cytokines, chemokines, and growth factors in the plasma samples: IL-1α, IL-1β, IFN-α2, IL-6, IL-12, IL-12 (p40), IL-18, LIF, MIF, TNF-α, TNF-β, TRAIL, IL-2, IFN-γ, IL-4, IL-5, IL-13, IL-10, IL-15, IL-16, IL-17, IL-1Rα, IL-2Rα, IL-3, IL-7, IL-9, BASIC-FGF, bNGF, PDGF-BB, G-CSF, HGF, SCF, M-CSF, SCGFb, MIP-1a, MIP-1b, MCP-1, MCP-3, IL-8, CTACK, RANTES, EOTAXIN, SDF1a, MIG, GROa, and IP-10. The cytokine levels were assessed by Luminex technology (Bio-Plex Workstation; Bio-Rad Laboratories, USA). The analysis of data was performed using software provided by the manufacturer (Bio-Rad Laboratories, USA).

The cell characterization was performed by Flow cytometry, including the cell populations: T lymphocytes, CD3^+^; CD4 T lymphocytes, CD3^+^CD4^+^; CD8 T lymphocytes, CD3^+^CD8^+^; T regulatory cells (T_REG_), CD3^+^CD4^+^CD25^+^CD127^-^FoxP3^+^; NK cells, CD3^-^CD56^+^; CD4 T effector memory cells (CD4 T_EM_), CD3^+^CD4^+^CD62L^-^CD95^+^CCR7^-^; CD8 T effector memory cells (CD8 T_EM_), CD3^+^CD8^+^CD62L^-^CD95^+^CCR7^-^; CD4 T central memory cells (CD4 T_CM_), CD3^+^CD4^+^CD62L^+^CD95^+^CCR7^+^; CD8 T effector memory cells (CD8 T_CM_) CD3^+^CD8^+^CD62L^+^CD95^+^CCR7^+^; CD4 T exhausted cells (CD4 T_EX_), CD3^+^CD4^+^PD-1^+^; CD8 T exhausted cells (CD8 T_EX_), CD3^+^CD8^+^PD-1^+^.

### Peripheral blood samples

We collected a total of 67 peripheral blood samples from patients with (n=40) or without cancer (n=27). Among cancer patients, 26 showed mild/moderate COVID-19 symptoms while 14 presented the severe form of the disease. Into the group of non-cancer patients, 12 were considered as mild/moderate COVID-19 group while 15 had the severe form. Briefly, peripheral blood was collected into EDTA tubes and plasma was separated after 15 min centrifugation for 400g and stored at -80°C for further cytokines analysis in Luminex. After that, blood was diluted in PBS (1:1) and mononuclear cells were isolated after Ficoll (Histopaque-1077, Sigma-Aldrich) density gradient centrifugation method (400g, 30min). PBMC was washed with PBS, followed by two washes with PBS + FBS 2%. Cell number was counted, and cell viability was measured by trypan blue exclusion method. Approximately, 5 x 10^6^ cells/vial were frozen in FBS with 2% DMSO in the liquid nitrogen storage tank. At an appropriate time, frozen PBMCs were thawed in a 37°C water bath, followed by two washes with 10mL of RPMI 1640 supplemented with 10% FBS and 1% L-glutamine. Cells were counted again using trypan blue dye for viability analysis, washed with PBS, and resuspended in PBS with 2% BSA for flow cytometry analysis.

### Flow cytometry analysis

Flow cytometry was used to characterize T cell composition in COVID-19 patients with or without cancer, focused on T cell activation/memory, exhaustion, and regulation. NK cell profile was also evaluated. For cell surface and intracellular staining, PBMCs were suspended in 20 µL of Fc block solution 2% in FACS buffer (PBS 1x, BSA1%) for 10 min. Then, for surface staining, cells were incubated for 30 min at room temperature using the following fluorochrome-conjugated anti-human antibodies from BD Biosciences: CD3-PerCP-Cy5.5(clone UCHT1), CD4-APC-H7 (clone RPA-T4), CD8-PE-Cy7 (clone RPA-T8), CD197 (CCR7)-BB515 (clone 2-L1-A), CD95-PE-Cy7 (clone DX2), CD62L-PE (clone DREG-56), CD279 (PD-1)-BB515 (EH12.1), CD25-PE-Cy7 (clone M-A251), CD127-Alexa Fluor 647 (clone HIL-7R-M21) and CD56-PE-Cy7 (clone B159). For FoxP3 intracellular staining (FoxP3-Alexa Fluor 488, clone 259D/C7 from BD Biosciences) we used the Transcription Factor Buffer Set (BD Biosciences) for fixation/permeabilization of cells, and protocol was performed as manufacturer instructions. Viable cells were identified by exclusion using Fixable Viability Stain 780 (BD Biosciences). Samples were analyzed using a BD FACS Canto II Cytometer (Becton Dickinson) and FlowJo software version 10.6.1 (TreeStar, San Diego, CA). Gating strategies were based on fluorescence-minus-one or negative controls.

### Statistical analysis

Statistics were performed using GraphPad Prism software version 7. Nonparametric one-way ANOVA with Kruskal-Wallis multiple test correction was used for statistical analysis of the results. Differences with *p* < 0.05 were considered statistically significant.

## Results

We analyzed a total of 67 patients diagnosed with COVID-19. Of these, 40 (59.7%) were cancer patients, and 27 (46.3%) were non-cancer patients. These groups were further divided into mild or severe cases of COVID-19, based on their need for mechanical ventilation and/or death due to the infection. Baseline characteristics are detailed in Table I. The median age in all four groups was higher than 45 years and the most frequent comorbidities were hypertension (41.6% - 64.3%) and diabetes (16.6%-42.1%) (Table I). Adenocarcinoma was the most common tumor subtype in cancer patients (30.1%), and stage IV was prevalent in mild and severe cases (31.5% and 57.1%, respectively) (Table I). Both groups had metastatic spread most frequently in the lungs, and 50% of severe cases were in palliative care. Mortality by COVID-19 reached 86% in severe cases in the cancer group, and 3.8% in mild cases, unrelated to the infection (Table I). No deaths were reported among non-cancer patients with mild COVID-19, but severe COVID-19 cases had a 66% mortality rate due to the infection (Table I).

**Table I:** Baseline characteristics of Cancer Patients with COVID-19.

|  | Non-Cancer Patients |  | Cancer Patients |  |
| --- | --- | --- | --- | --- |
|  | Mild COVID-19 | Severe COVID-19 | Mild COVID-19 | Severe COVID-19 |
| **N of patients** | 12 | 15 | 26 | 14 |
| **Gender M/F (%)** | 50/50 | 53/47 | 30/70 | 36/64 |
| **Median age (range)** | 58 (33-80) | 47 (26-93) | 58.5 (18-78) | 62 (29-78) |
| **Comorbidities** | Diabetes: 16.6%<br>HT: 41.6%<br>Obesity: 41.6%<br>Others: 16.6% | Diabetes: 46.6%<br>HT: 46.6%<br>Obesity: 6.6%<br>CF/CMP: 13.3% | Diabetes: 26.9%<br>HT: 57.7%<br>Obesity: 11%<br>CF/CMP: 7.7%<br>Others: 15.3% | Diabetes: 21.4%<br>HT: 64.3%<br>Obesity: 7.1%<br>CF/CMP: 21.4%<br>Others: 35.7% |
| **Tumor subtype** | N/A | N/A | AdenoCA: 26%<br>SCC: 7.6%<br>Sarcoma: 3.8%<br>Others: 54%<br>N/A: 8.6% | AdenoCA: 36%<br>SCC: 21%<br>Others: 36%<br>N/A: 7% |
| **Stage** | N/A | N/A | I: 3.8%<br>II: 7.7%<br>III: 19.2%<br>IV: 38.5%<br>N/A: 30.8% | I: 7.1%<br>II: 14.2%<br>IV: 57.1%<br>Non-mtx: 7.1%<br>N/A: 14.5% |
| **Metastatic sites** | N/A | N/A | Lungs: 11.5%<br>Liver: 3.85%<br>CNS: 3.5%<br>Bones: 15.4%<br>Lymph nodes: 3.9%<br>Skin: 7.7 % | Lungs: 21.4%<br>Liver: 14.3%<br>CNS: 14.3%<br>Bones: 7.14%<br>Lymph nodes: 14.3%<br>Peritoneum: 7.14% |
| **Intention to treat** | N/A | N/A | Adjuvant: 7.7%<br>Neoadjuvant: 15.4%<br>Palliative: 19.2%<br>Curative: 19.2%<br>Control: 15.4%<br>Support: 19.2% | Adjuvant: 4.14%<br>Palliative: 50%<br>Curative: 7.14%<br>Control: 21.4%<br>Support: 21.4% |
| **Deaths; Cause** | N/A | 66%; COVID-19 | 3.8%; unrelated to COVID-19 | 86%; COVID-19 |

N: number; F: female; M: male; HT: hypertension; CF: cardiac failure; CMP: cardiomyopathies; AdenoCA: adenocarcinoma; SCC: squamous cell carcinoma; Non-mtx: non-metastatic; CNS: central nervous system; N/A: not applicable.

Regarding the T cell characterization, cancer patients with mild COVID-19 presented increased T CD3^+^ population compared to cancer patients with severe COVID-19 (Figure 1A). In addition, considering exclusively COVID-19 severe cases, the T CD8^+^ population was higher in cancer patients compared to the non-cancer group (Figure 1A). T CD4^+^ lymphocytes proportions showed no significant differences between our participants (Figure 1A).

**Figure 1:**
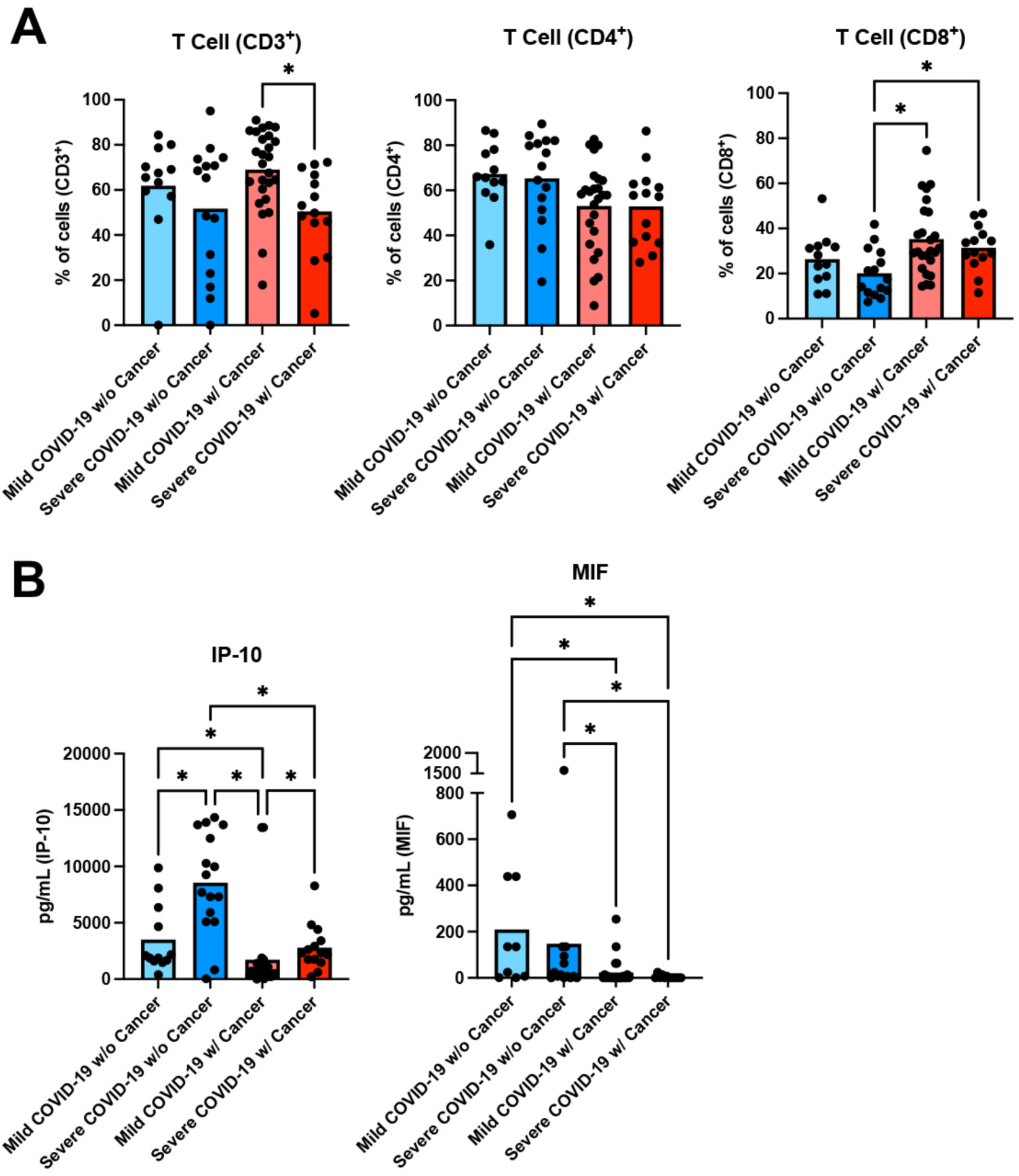
Analysis of T lymphocytes and cytokines from peripheral blood samples. Blood samples were analyzed from mild and severe COVID-19 hospitalized patients with cancer and without cancer. Each individual dot represents one patient and collum represents the mean. (A) T lymphocytes analysis in each of the participants were done by flow cytometry as CD3^+^, CD3^+^CD4^+^ or CD3^+^CD8^+^ lymphocytes. (B) Cytokines (CXCL10/IP-10 and MIF) analysis in each of the participants were done by Luminex. Nonparametric of one-way ANOVA with multiple comparisons and two-stage step-up method of Benjamini, Krieger and Yekutieli were used for statistical analysis of the results. (*) mean *p*<0.05 between selected groups.

Analysis of cytokine levels in plasma demonstrated that the chemokine CXCL-10 (IP-10) was increased in cancer patients compared to non-cancer patients with COVID-19, both in mild cases and severe cases (Figure 1B). Comparing cancer patients, mild COVID-19 patients exhibited lower CXCL-10 levels than severe COVID-19 patients (Figure 2B). Furthermore, MIF was significantly lower in cancer patients with COVID-19 compared to non-cancer patients (Figure 1B). However, our results showed no significant differences between cancer patients with non-cancer patients in all other cytokines analysis (data not shown).

**Figure 2:**
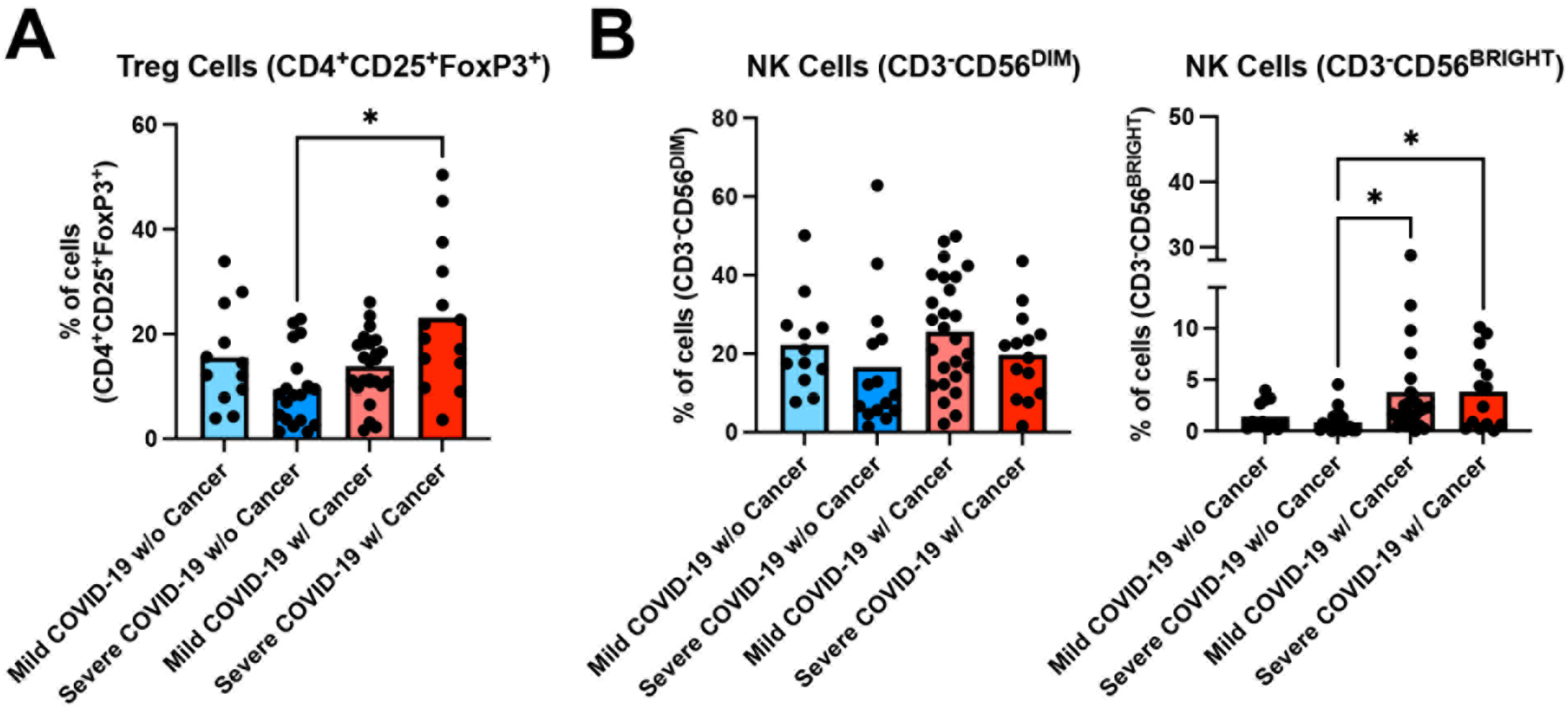
Analysis of Treg and NK cells from peripheral blood samples. Blood samples were analyzed from mild and severe COVID-19 hospitalized patients with cancer and without cancer. Each individual dot represents one patient and collum represents the mean. (A) Treg lymphocytes analysis in each of the participants were done by flow cytometry as CD3^+^CD4^+^CD25^+^CD127^-^ FoxP3^+^ or (B) NK cells as CD3^-^CD56^+^. Nonparametric of one-way ANOVA with multiple comparisons and two-stage step-up method of Benjamini, Krieger and Yekutieli were used for statistical analysis of the results. (*) mean *p*<0.05 between selected groups.

We also analyzed different subsets of T regulatory (Treg) cells and NK cells (Figure 2). Considering severe COVID-19 cases, cancer patients presented higher proportions of Treg cells (CD4^+^CD25^+^FoxP3^+^) than non-cancer patients (Figure 2A). No differences were observed for NK cell lineages CD3^+^CD56^DIM^ (Figure 2B). However, the CD3^-^CD56^BRIGHT^ population was significantly elevated in cancer patients with severe COVID-19 compared to non-cancer cases (Figure 2B). Furthermore, memory T cells analysis (central and effector memory) revealed that cancer patients with COVID-19 exhibited lower levels of effector memory and higher levels of central memory cells relative to non-cancer patients (Figure 3).

**Figure 3:**
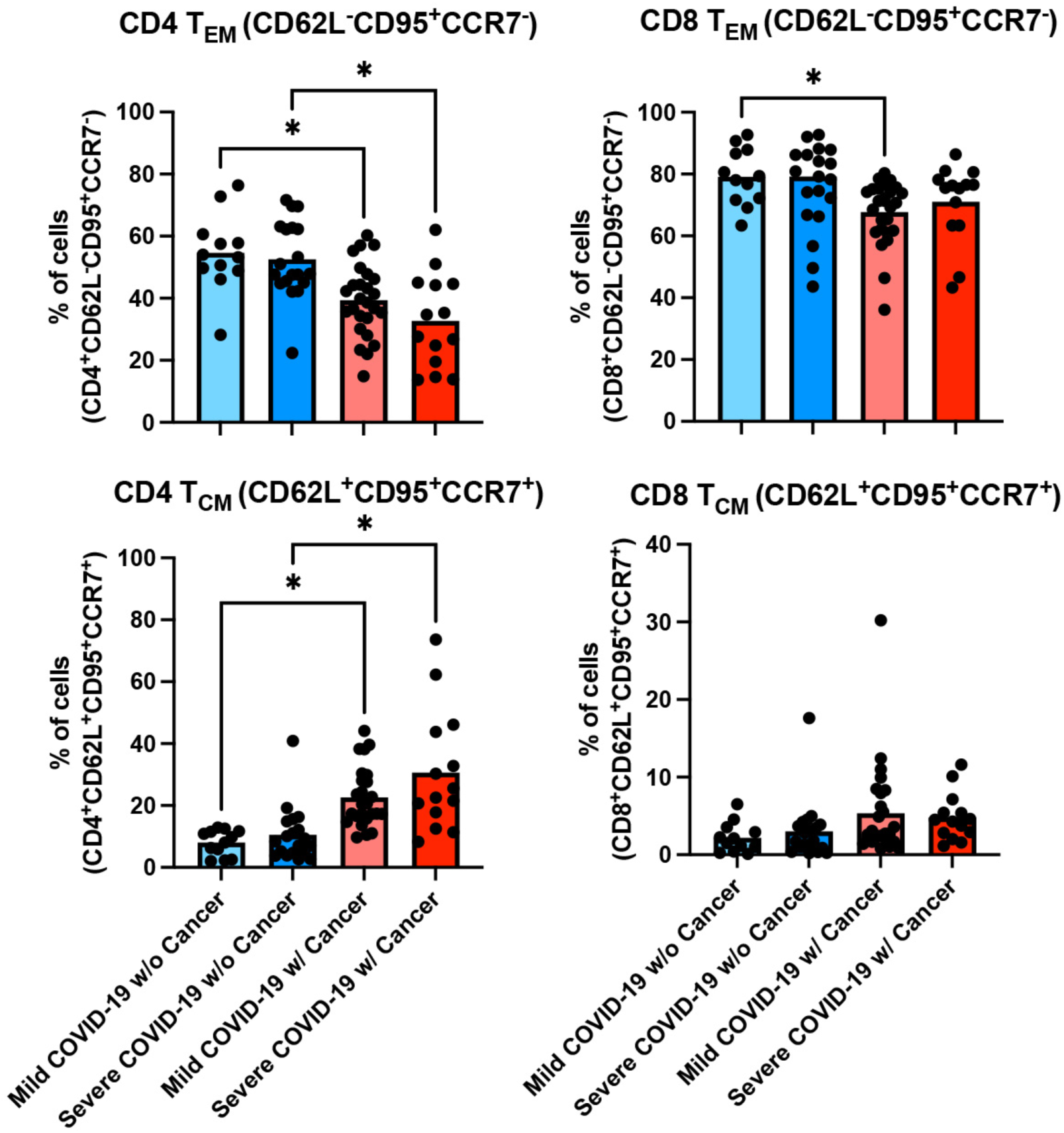
Analysis of memory cells from peripheral blood samples. Blood samples were analyzed from mild and severe COVID-19 hospitalized patients with cancer and without cancer. Each individual dot represents one patient and collum represents the mean. Effector memory (EM) T lymphocytes analysis in each of the participants were done by flow cytometry as CD3^+^CD4^+^CD62L^-^CD95^+^CCR7^-^ (CD4 T_CM_) or CD3^+^CD8^+^CD62L^-^CD95^+^CCR7^-^ (CD8 T_CM_). Central memory (CM) T lymphocytes analysis in each of the participants were done by flow cytometry as CD3^+^CD4^+^CD62L^+^CD95^+^CCR7^+^ (CD4 T_CM_) or CD3^+^CD8^+^CD62L^+^CD95^+^CCR7^+^ (CD8 T_CM_). Nonparametric of one-way ANOVA with multiple comparisons and two-stage step-up method of Benjamini, Krieger and Yekutieli were used for statistical analysis of the results. (*) mean *p*<0.05 between selected groups.

Finally, we analyzed the T cell exhaustion profile in cancer and non-cancer patients (Figure 4). Among mild COVID-19 cases, cancer patients showed increased levels of exhausted CD4 (CD4^+^PD-1^+^) and CD8 T (CD8^+^PD-1^+^) cells compared with non-cancer patients (Figure 4). Similarly, among severe COVID-19 cases, cancer patients also showed increased levels of exhausted T cells (CD4^+^PD-1^+^ and CD8^+^PD-1^+^) compared to non-cancer patients (Figure 4).

**Figure 4:**
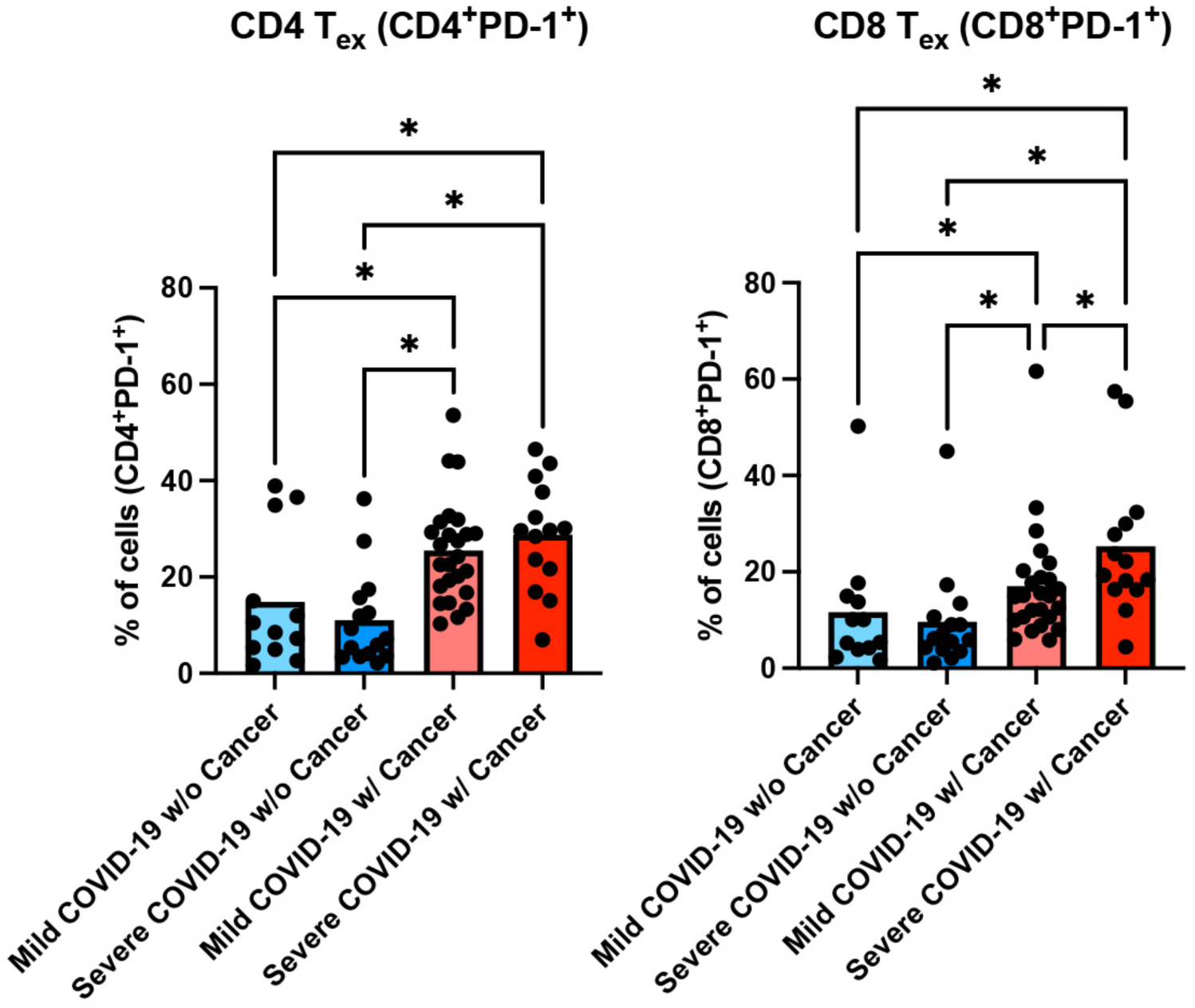
Analysis of exhausted T lymphocyte from peripheral blood samples. Blood samples were analyzed from mild and severe COVID-19 hospitalized patients with cancer and without cancer. Each individual dot represents one patient and collum represents the mean. Exhausted (EX) T lymphocytes analysis in each of the participants were done by flow cytometry as CD3^+^CD4^+^PD-1^+^ (CD4 T_EX_) or CD3^+^CD8^+^PD-1^+^ (CD8 T_EX_). Nonparametric of one-way ANOVA with multiple comparisons and two-stage step-up method of Benjamini, Krieger and Yekutieli were used for statistical analysis of the results. (*) mean *p*<0.05 between selected groups.

## Discussion

The population with comorbidities infected with COVID-19 faces higher mortality rates and poorer outcomes, including longer hospital stays and intubation requirements (11). This impact is particularly significant in cancer patients, with an estimated mortality rate of 6% in 2020 (12). Despite attempts to explain these worsened outcomes by attributing them to cancer patients’ immunosuppression and treatments, underlying factors in oncologic patients, such as age, smoking status, hypertension, chronic lung disease, and coronary artery disease were observed as more likely to increase their mortality with severe COVID-19 than the cancer itself (11, 13,14). Ongoing research on cancer patients with COVID-19 aims their better management by adjusting current immunosuppressive treatments, using targeted therapies, and effectively indicating hospitalization (15,16). Therefore, cytokines and lymphocyte activation profiles are investigated to enhance our comprehension of the immune response’s role in the outcomes of the infection in Oncology (15,16,17).

The chemokine CXCL-10 (IP-10) plays a role in the initial cascade of the cytokine storm in inflammatory states, although their correlation with both cancer and COVID-19 severity are not fully elucidated [19]. Our study found a positive association of CXCL-10 with disease severity in COVID-19 cases (Figure 1B). Interestingly, we also observed a negative association between CXCL-10 and MIF levels and the presence of cancer (Figure 1B). Therefore, it may serve as a potential biomarker for assessing the severity of the infection, which supports the use of targeted therapies against CXCL-10 in severe COVID-19 cases, especially in cancer patients (19).

Beyond cytokines, the lymphocyte activation profile is believed to be involved in COVID-19 severity among cancer patients, particularly with the use of immunotherapy and immunosuppressive therapies (13,15). While some studies suggest worsened outcomes with these agents, a recent cohort study involving cancer patients concluded that they were not solely responsible for the severity of COVID-19 or the development of cytokine storms, supporting their continuation (15).

Notably, COVID-19 was previously associated with lymphopenia, impairing a prompt immune response against the infection, primarily coordinated by T cells (15). Thibaudin *et al.* (2020) demonstrated an accentuated depletion of T CD3^+^ cells in cancer patients with the infection (20). Consistent with our analysis, which showed a significant association between T CD3^+^ depletion and the severity of COVID-19 (Figure 1), this could potentially hinder an appropriate T cell response. Although severe COVID-19 patients exhibited higher percentages of T CD8^+^ cells, this cell population could present impaired function, marked by increased exhaustion markers and prominent T regulatory cells within this subset. Notably, our results demonstrated an increased population of T regulatory cells in cancer patients with severe COVID-19 compared to non-cancer patients (Figure 2). Therefore, this suggests a suppressed T cell activation against COVID-19 infection and increased regulatory activity, potentially impairing the antiviral activity of lymphocytes and immunoglobulins release against the viral infection (21,22).

Additionally, NK cells are involved in the innate system response against viral infections, while also influencing the tumor microenvironment (25). Regarding COVID-19, previous research indicated that severe cases were associated with a decreased proportion and effector function of NK cells, potentially impairing viral clearance and immune regulation against inflammatory mediators. Our results showed increased rates of CD56^BRIGHT^ NK cells in cancer patients compared to non-cancer patients with COVID-19 (Figure 2). Although this could suggest an increased dysregulation in cancer patients, the precise role of these cells in COVID-19 is still uncertain. Therefore, further investigations are needed for a clear understanding of their antiviral activity in COVID-19 as well as their impact on oncologic patients (25,26,27).

The exhaustion pattern of T cells is concerning in the tumoral microenvironment, as it results in a loss of effective immunological responses to tumor antigens, further increasing its spreading capacity (21,22). Recent studies correlated COVID-19 with T lymphocyte exhaustion, characterized by elevated levels of PD-1 and Tim-3 in CD8^+^ and CD4^+^ lymphocytes (11,15). Given the high expression of PD-1 ligands in cancer patients, this additional feature of COVID-19 infection could augment the exhaustive pattern, facilitate the tumoral escape from host defenses, and worsen their outcomes. Consistent with existing literature, our findings indicate a positive correlation between cancer and the severity of COVID-19 with increased levels of PD-1 (Figure 4), supporting the potential use of PD-1 as a predictive biomarker for individualized management (15). Furthermore, in line with this hypothesis, the investigation of anti-PD-1 targeted therapies is currently underway in clinical trials as a potential management strategy (23,24).

This study has several limitations. The relatively small number of patients may limit the generalizability of the findings. Since the study was conducted at a single center, the results may not be representative of broader populations. Additionally, all patients were analyzed during the early stages of the COVID-19 pandemic, prior to the availability of vaccinations, which could influence disease outcomes and management strategies. These factors should be considered when interpreting the results, and further multicenter studies with larger cohorts in the post-vaccination era are warranted to validate our findings.

Our results highlight the importance of assessing the lymphocyte activation profile in cancer patients with COVID-19. Understanding the alterations in specific T cell subsets can provide insights into the immune response and potential immune dysregulation in this population. However, our study has limitations. Taken together, our findings suggest that COVID-19 patients with cancer exhibit amplification of regulatory lymphocytes, including T and NK cells, along with an exhausted immune profile, which could potentially impact cancer clinical outcomes. Furthermore, COVID-19 cancer patients exhibited a shift from effector memory to central memory T-cells. Additionally, we observed a positive association between CXCL-10 (IP-10) levels and disease severity in COVID-19 cases. These findings provide valuable insights into the immunological mechanisms underlying clinical outcomes and enhance the identification of potential biomarkers for severe disease, which could be targeted on therapies aiming to mitigate adverse clinical outcomes.

## Acknowledgements

We thank the INCA Flow Cytometry Core Facility for technical support with FACS analysis and the FIOCRUZ Luminex Core Facility for technical support with Luminex data analysis.

## Funding

This work was supported by grants to JPBV from CNPq (311926/2021-5) and FAPERJ (210.168/2020, 211.126/2021 and 200.568/2023). MMB was supported by CNPq fellowship; BCP was supported by a CAPES fellowship, and LRG was supported by a FAPERJ fellowship.

## Availability of data and materials

The data generated in the present study may be requested from the corresponding author.

## Authors’ contributions

PTB and JPBV designed research; KLS, BCP, LRG, IA-Q and EDH processed patient samples; MMB, BCP, KLS, IA-Q and EDH performed and analyzed the FACS experiments; MMB, BCP, IA-Q and EDH performed and analyzed the Luminex experiments; FAB, MAS, ACM, EDH, PTB and JPBV participated in clinical data analysis; MMB, PTB and JPBV wrote the paper. All authors have read and approved the final manuscript.

## Patient consent for publication

Not applicable.

## Competing interests

The authors declare that they have no competing interests.

## Notes

### Competing Interest Statement

The authors have declared no competing interest.

## References

1. World Health Organization. WHO-convened global study of origins of SARS-CoV-2: China Part [Internet]. Geneva: World Health Organization; 2021 [cited 2025 Apr 17]. Available from: https://www.who.int/publications/i/item/who-convened-global-study-of-origins-of-sars-cov-2-china-part

2. de Azambuja E, Brandao M, Wildiers H, Laenen A, Aspeslagh S, Fontaine C, Collignon J, Lybaert W, Verheezen J, Rutten A, et al: Impact of solid cancer on in-hospital mortality overall and among different subgroups of patients with COVID-19: a nationwide, population-based analysis. ESMO Open 5:e000947, 2020.

3. Fernandes Q, Inchakalody VP, Merhi M, Mestiri S, Taib N, Moustafa Abo El-Ella D, Bedhiafi T, Raza A, Al-Zaidan L, Mohsen MO, et al: Emerging COVID-19 variants and their impact on SARS-CoV-2 diagnosis, therapeutics and vaccines. Ann Med 54:524–540, 2022.

4. Negahdaripour M, Shafiekhani M, Moezzi SMI, Amiri S, Rasekh S, Bagheri A, Mosaddeghi P, Vazin A: Administration of COVID-19 vaccines in immunocompromised patients. Int Immunopharmacol 99:108021, 2021.

5. Fendler A, de Vries EGE, GeurtsvanKessel CH, Haanen JB, Wörmann B, Turajlic S, von Lilienfeld-Toal M: COVID-19 vaccines in patients with cancer: immunogenicity, efficacy and safety. Nat Rev Clin Oncol 19:385–401, 2022.

6. Larson C, Oronsky B, Goyal S, Ray C, Hedjran F, Hammond TC, et al. COVID-19 and cancer: A guide with suggested COVID-19 rule-out criteria to support clinical decision-making. Biochimica et Biophysica Acta (BBA) - Reviews on Cancer. 2020 Dec;1874(2):188412.

7. Li M, Guo W, Dong Y, Wang X, Dai D, Liu X, Wu Y, Li M, Zhang W, Zhou H, et al: Elevated exhaustion levels of NK and CD8+ T cells as indicators for progression and prognosis of COVID-19 disease. Front Immunol 11:580237, 2020.

8. Kandeel EZ, Refaat L, Bayoumi A, Nooh HA, Hammad R, Khafagy M, Abdellateif MS: The role of lymphocyte subsets, PD-1, and FAS (CD95) in COVID-19 cancer patients. Viral Immunol 35:491–502, 2022.

9. World Health Organization: Clinical management of severe acute respiratory infection when COVID-19 is suspected. 2020 May 27. Available from: https://www.who.int/publications-detail/clinical-management-of-severe-acute-respiratory-infection-when-novel-coronavirus-(ncov)-infection-is-suspected

10. Centers for Disease Control and Prevention: CDC 2019-Novel Coronavirus (2019-nCoV) Real-Time RT-PCR Diagnostic Panel. 2020 Jul 13. Available from: https://www.fda.gov/media/134922/download

11. Pathania AS, Prathipati P, Abdul BA, Chava S, Katta SS, Gupta SC, Gangula PR, Pandey MK, Durden DL, Byrareddy SN, Challagundla KB: COVID-19 and cancer comorbidity: therapeutic opportunities and challenges. Theranostics 11:731–753, 2021.

12. Wu Z and McGoogan JM: Characteristics of and important lessons from the coronavirus disease 2019 (COVID-19) outbreak in China: summary of a report of 72 314 cases from the Chinese Center for Disease Control and Prevention. JAMA 323:1239–1242, 2020.

13. Aboueshia M, Hussein MH, Attia AS, Swinford A, Miller P, Omar M, Toraih EA, Saba N, Safah H, Duchesne J, et al: Cancer and COVID-19: analysis of patient outcomes. Future Oncol 17:3499–3510, 2021.

14. Zarifkar P, Kamath A, Robinson C, Morgulchik N, Shah SFH, Cheng TKM, Dominic C, Fehintola AO, Bhalla G, Ahillan T, et al: Clinical characteristics and outcomes in patients with COVID-19 and cancer: a systematic review and meta-analysis. Clin Oncol (R Coll Radiol) 33:e180–e191, 2021.

15. Bakouny Z, Labaki C, Grover P, Awosika J, Gulati S, Hsu CY, Alimohamed SI, Bashir B, Berg S, Bilen MA, et al: Interplay of immunosuppression and immunotherapy among patients with cancer and COVID-19. JAMA Oncol 9:128–134, 2023.

16. Addeo A, Obeid M and Friedlaender A: COVID-19 and lung cancer: risks, mechanisms and treatment interactions. J Immunother Cancer 8:e000892, 2020.

17. Pignataro-Oshiro F, Figueiredo AB, Galdino NAL, Morais KLP, Dutra WO, Silva BGM, Feriani D, Abrantes FA, Silva ILAFE, Filho JS, et al: Distinct systemic immune networks define severe vs. mild COVID-19 in hematologic and solid cancer patients. Front Immunol 13:1052104, 2023.

18. Hirano T: IL-6 in inflammation, autoimmunity and cancer. Int Immunol 33: 127–148, 2021.

19. Coperchini F, Chiovato L and Rotondi M: Interleukin-6, CXCL10 and infiltrating macrophages in COVID-19-related cytokine storm: not one for all but all for one. Front Immunol 12:668507, 2021.

20. Thibaudin M, Fumet JD, Bon M, Hampe L, Limagne E and Ghiringhelli F: Immunological features of coronavirus disease 2019 in patients with cancer. Eur J Cancer 139:70–80, 2020.

21. Møller SH, Hsueh PC, Yu YR, Zhang L and Ho PC: Metabolic programs tailor T cell immunity in viral infection, cancer, and aging. Cell Metab 34:378–395, 2022.

22. Liu X, Si F, Bagley D, Ma F, Zhang Y, Tao Y, Shaw E and Peng G: Blockades of effector T cell senescence and exhaustion synergistically enhance antitumor immunity and immunotherapy. J Immunother Cancer 10:e005020, 2022.

23. ClinicalTrials.gov [Internet]: National Library of Medicine (US). Identifier NCT04413838, Efficiency and security of nivolumab therapy in obese individuals with COVID-19 (COrona Virus Disease) infection. 2020 Jun 04 [cited 2024 Feb 13]. Available from: https://clinicaltrials.gov/study/NCT04413838.

24. ClinicalTrials.gov [Internet]: National Library of Medicine (US). Identifier NCT04268537, Immunoregulatory therapy for 2019-nCoV. 2020 Feb 13 [cited 2024 Feb 13]. Available from: https://clinicaltrials.gov/study/NCT04268537.

25. Santoni G, Amantini C, Santoni M, Maggi F, Morelli MB and Santoni A: Mechanosensation and mechanotransduction in natural killer cells. Front Immunol 12:688918, 2021.

26. van Eeden C, Khan L, Osman MS and Cohen Tervaert JW: Natural killer cell dysfunction and its role in COVID-19. Int J Mol Sci 21:6351, 2020.

27. Manickam C, Sugawara S and Reeves RK: Friends or foes? The knowns and unknowns of natural killer cell biology in COVID-19 and other coronaviruses in July 2020. PLoS Pathog 16:e1008820, 2020.

